# Direct Economic Burden of Mental Health Disorders Associated with Polycystic Ovary Syndrome: Systematic Review and Meta-analysis

**DOI:** 10.1101/2023.01.05.23284220

**Authors:** Surabhi Yadav, Olivia Delau, Adam Bonner, Daniela Markovic, William Patterson, Sasha Ottey, Richard P. Buyalos, Ricardo Azziz

**Affiliations:** School of Global Public Health, New York University, New York, NY 10003, USA; Department of Obstetrics & Gynecology, Heersink School of Medicine, University of Alabama at Birmingham, Birmingham, AL 35294, USA; Division of General Internal Medicine and Health Services Research, UCLA, Los Angeles, CA 90095, USA; PCOS Challenge: The National Polycystic Ovary Syndrome, Atlanta, GA 30308, USA; Department of Obstetrics and Gynecology, UCLA, Los Angeles, CA 90095, USA; Department of Medicine, Heersink School of Medicine, University of Alabama at Birmingham, Birmingham, AL 35294, USA; Department of Health Policy, Management and Behavior, School of Public Health, University at Albany, SUNY, Rensselaer, NY 12144, USA; Department of Healthcare Organization and Policy, School of Public Health, University of Alabama at Birmingham, Birmingham, AL 35294, USA

**Keywords:** polycystic ovarian syndrome, economic burden, mental health disorders, anxiety, depression, eating disorder, postpartum depression

## Abstract

**BACKGROUND:** Polycystic ovary syndrome (PCOS) is the most common hormone disorder affecting about one in seven reproductive-aged women worldwide and approximately 6 million women in the United States (U.S.). PCOS can be a significant burden to those affected and is associated with an increased prevalence of mental health (MH) disorders such as depression, anxiety, eating disorders, and postpartum depression. We undertook this study to determine the excess economic burden associated with MH disorders in women with PCOS, in order to allow for a more accurate prioritization of the disorder as a public health priority.

**METHODS:** Followed PRISMA reporting guidelines for systematic review, we searched PubMed, Web of Science, EBSCO, Medline, Scopus, and PsycINFO through July 16, 2021, for studies on MH disorders in PCOS. Excluded were studies not in humans, without controls, without original data, or not peer reviewed. As anxiety, depression, eating disorders, and postpartum depression were by far the most common MH disorders assessed by the studies, we performed our meta-analysis on these disorders. Meta-analyses were performed using the DerSimonian-Laird random-effects model to compute pooled estimates of prevalence ratios (PR) for the associations between PCOS and these MH disorders, and then calculated the excess direct costs of related to these disorders in U.S. dollars (USD) for women suffering from PCOS in the U.S. alone. The quality of selected studies was assessed using the Newcastle-Ottawa Scale.

**RESULTS:** We screened 78 articles by title/abstract, assessed 43 articles in full-text, and included 25 articles. Pooled PRs were 1.42 (95% CI: 1.32-1.52) for anxiety, 1.65 (95% CI: 1.44-1.89;) for depression, 1.48 (95% CI: PR: 1.06-2.05) for eating disorders, and 1.20 (95% CI: 0.96-1.50) for postpartum depression, for PCOS relative to controls. In the U.S, the additional direct healthcare costs associated with anxiety, depression and eating disorders in PCOS were estimated to be $1.939 billion/yr., 1.678 billion/yr., and $0.644 billion/yr. in 2021 USD, respectively. Postpartum depression was excluded from the cost analyses due to the non-significant meta-analysis result. Taken together, the additional direct healthcare costs associated with anxiety, depression and eating disorders in PCOS was estimated to be $4.261 billion/yr. in 2021 USD.

**CONCLUSIONS:** Overall, the direct healthcare annual costs for the most common MH disorders in PCOS, namely anxiety, depression, and eating disorders exceeds $4 billion in 2021 USD for the U.S. population alone. Taken together with our prior work, these data suggest that the healthcare-related economic burden of PCOS exceeds $15 billion yearly, considering the costs of PCOS diagnosis, and cost related to PCOS-associated MH, reproductive, vascular, and metabolic disorders. As PCOS has much the same prevalence across the world, the excess economic burden attributable to PCOS globally is enormous, mandating that the scientific and policy community increase its focus on this important disorder.

**FUNDING:** The study was supported, in part, by PCOS Challenge: The National Polycystic Ovary Syndrome Association and by the Foundation for Research and Education Excellence

**CLINICAL TRIAL NUMBER:** N/A

## INTRODUCTION

Polycystic ovary syndrome (PCOS) is a highly prevalent disorder, highly inherited complex polygenic, multifactorial disorder (1). PCOS single most common endocrine-metabolic disorder in reproductive-aged women today, affecting 5-15% of unselected reproductive-aged women (1990 National Institutes of Health criteria), and potentially represents a significant financial burden to our health care (2,3). Pathophysiological abnormalities in gonadotropin secretion or action, ovarian folliculogenesis, steroidogenesis, insulin secretion or action, and adipose tissue function, among others, have been described in PCOS. Women with PCOS are at increased risk for glucose intolerance and type 2 diabetes mellitus (T2DM), hepatic steatosis and metabolic syndrome, hypertension, dyslipidemia, vascular thrombosis, cerebrovascular accidents, possibly cardiovascular events, subfertility and obstetric complications, endometrial carcinoma, and mood and psychosexual disorders (1).

Although the physical symptoms of PCOS are increasingly recognized by practicing clinicians, little attention has focused on the psychological correlates of this frequent endocrine disorder (4,5). A significant amount of research has been conducted showing the direct causal relationship between PCOS diagnosis and mental health (MH) disorders. PCOS is associated with an increased risk of a diagnosis of depression, anxiety, bipolar disorder, and obsessive-compulsive disorder (OCD), and is associated with worse symptoms of depression, anxiety, OCD, and somatization (4,5). Screening for these disorders to allow early intervention may be warranted.

In order to allow for a more accurate prioritization of the disorder as a public health priority, we have pursued a comprehensive estimation of the economic burden of PCOS. In previous studies conducted by our team, we estimated the mean annual cost of the initial evaluation of PCOS to be $93 million, that of hormonally treating menstrual dysfunction/abnormal uterine bleeding to be $1.35 billion, that of providing infertility care to be $533 million, and that of treating hirsutism to be $622 million in 2014 USD (2). In a more recent study, we estimated the costs of PCOS-associated T2DM and associated stroke to be $1.5 billion and $2.4 billion in 2020 USD, respectively (3). The present study aims to assess the direct healthcare-related economic burden of PCOS-related MH disorders. To do so, we conducted a systematic review and meta-analysis of published studies with human subjects and controls that analyzed the relationship between MH disorders and previous diagnosis of PCOS, and then calculated the related direct economic burden.

## MATERIALS & METHODS

### Systematic Review

A systematic review was performed, adhering to the Preferred Reporting Items for Systematic Reviews and Meta-analyses (PRISMA) Statement and Checklist (https://www.prisma-statement.org/), for reports examining the relationship of MH disorders and PCOS through July 16, 2021. The systematic review was conducted on the six English databases (i.e., PubMed, Web of Science, EBSCO, Medline, Scopus, and PsycINFO). The following keywords were used (mental health OR mental illness OR mental disorder OR psych* OR anxiety OR depression OR quality of life OR eating disorder OR bulimia OR postpartum depression) AND (‘cost* OR ‘economic burden’ OR ‘cost-of-illness OR ‘burden of illness’), “Depressive Disorder/economics”[MAJR], “PCOS” AND “economic burden” OR “costs” OR “cost-of-illness” OR “burden of illness”, “PCOS” AND “economic burden” AND “mental health”, ‘polycystic ovary syndrome’ AND ‘anxiety, “Polycystic Ovary Syndrome/psychology”[MAJR] (**Supplemental table 1**) (6).

### Study eligibility criteria

Studies were eligible for inclusion if they: (a) were original peer-reviewed academic articles; AND (b) were observational studies that presented accurate and precise data regarding the risk, including reporting relative risks, odds ratios, hazard ratios, or prevalence or incidence rates, of MH disorders in women with PCOS compared with a control group. MH disorders included depressive disorders, such as major depression disorder, dysthymia, minor depression or subclinical/subthreshold depression, or affective disorders containing depressive disorders, emotional distress, eating disorders, or mood and anxiety disorders. In the case of repeatedly published and studied literature based on the same batch of data or sample population the most recent studies with the complete data set were included. Studies were excluded if they: (a) were based on non-human species; (b) did not have full text available; (c) did not include a control group; (d) reported solely on diseases other than PCOS; or (e) were reviews, letters, or commentaries.

### Study selection and data extraction

Search strategy and study identification was performed by one investigator (S.Y.) using a standardized approach. Articles selected for inclusion were then screened by four authors (O.D., A.B., S.Y., D.M.). Further, four investigators worked to independently extract data on study characteristics and outcomes (A.B., S.Y., O.D., D.M.). Disagreements were discussed until consensus was reached.

### Quality assessment

Four investigators worked in duplicate to independently assess the quality of eligible studies using the Newcastle-Ottawa scale (S.Y., O.D., A.B., R.A.) (https://www.ohri.ca/programs/clinical_epidemiology/oxford.asp).

### Statistical analysis

Following the systematic search, the data were submitted to a meta-analysis to estimate the degree of relationship between PCOS and four dichotomous outcomes including anxiety, depression, eating disorders and postpartum depression. The meta-analysis was performed using the DerSimonian-Laird random effects model (7) and the results of the analyses were summarized using the pooled PR and its 95% confidence intervals for each of the above outcomes.

We should note that when estimating risk, in general we can assume that the odds ratio (OR) approximates the risk ratio when the prevalence in both groups is low (∼ <10%). However, when the prevalence of the outcome is higher the OR is likely to over-estimate of risk ratio. Outcomes for this study were generally higher so we chose to use prevalence ratios (PRs) instead of odds ratios (ORs) for the economic burden calculations. The limitation of using PRs is that these effects were not adjusted for the full set of covariates as for the OR analysis. However, most studies used groups that were matched by age and/or BMI by design.

Study specific and overall effect estimates were visually presented using Forest plots. Between-study heterogeneity was evaluated using the I2 statistic. In case of significant heterogeneity (I2 > 70%) sensitivity analyses were performed by excluding any outliers from the analysis. We defined an outlier as any study whose confidence intervals did not overlap the confidence interval of the pooled estimate for the purpose of the sensitivity analysis. Publication bias was assessed using funnel plots. Additionally, to adjust for possible publication bias, we recalculated the results using the ‘trim and fill’ method (8). Analyses were performed using R version 4.1.3.

As previously (2,3), we defined the prevalence of PCOS based on the NIH criteria (phenotypes A and B of the Rotterdam criteria) or what is considered the ‘classic’ PCOS phenotypes), which we have conservatively estimated to be 6.6%. Annual cost data for medical treatment of depression for individuals 18-49 years of age was obtained from a 2021 study (9), for anxiety for ages 15-54 was obtained from a 1999 study (10), and for eating disorders for individuals 20-49 years of age was obtained from a 2021 study (11). We adjusted for inflation using the medical care inflation calculator (https://www.officialdata.org/Medical-care/price-inflation/).

## RESULTS

A total of 2018 studies were identified during the initial literature review, of which 78 were screened by title and abstract (**Fig. 1**). Forty-three potentially eligible studies were reviewed in detail, of which 18 were excluded due to insufficient information. For example, some studies did not include information on a control group, did not include measures of association, used a continuous outcome instead of dichotomous outcome, or did not provide information about risk/prevalence that was needed to compute the prevalence ratios. The general characteristics of the 25 included studies (12-36) are detailed in **Supplemental Tables 2-5**.

**Fig 1.**
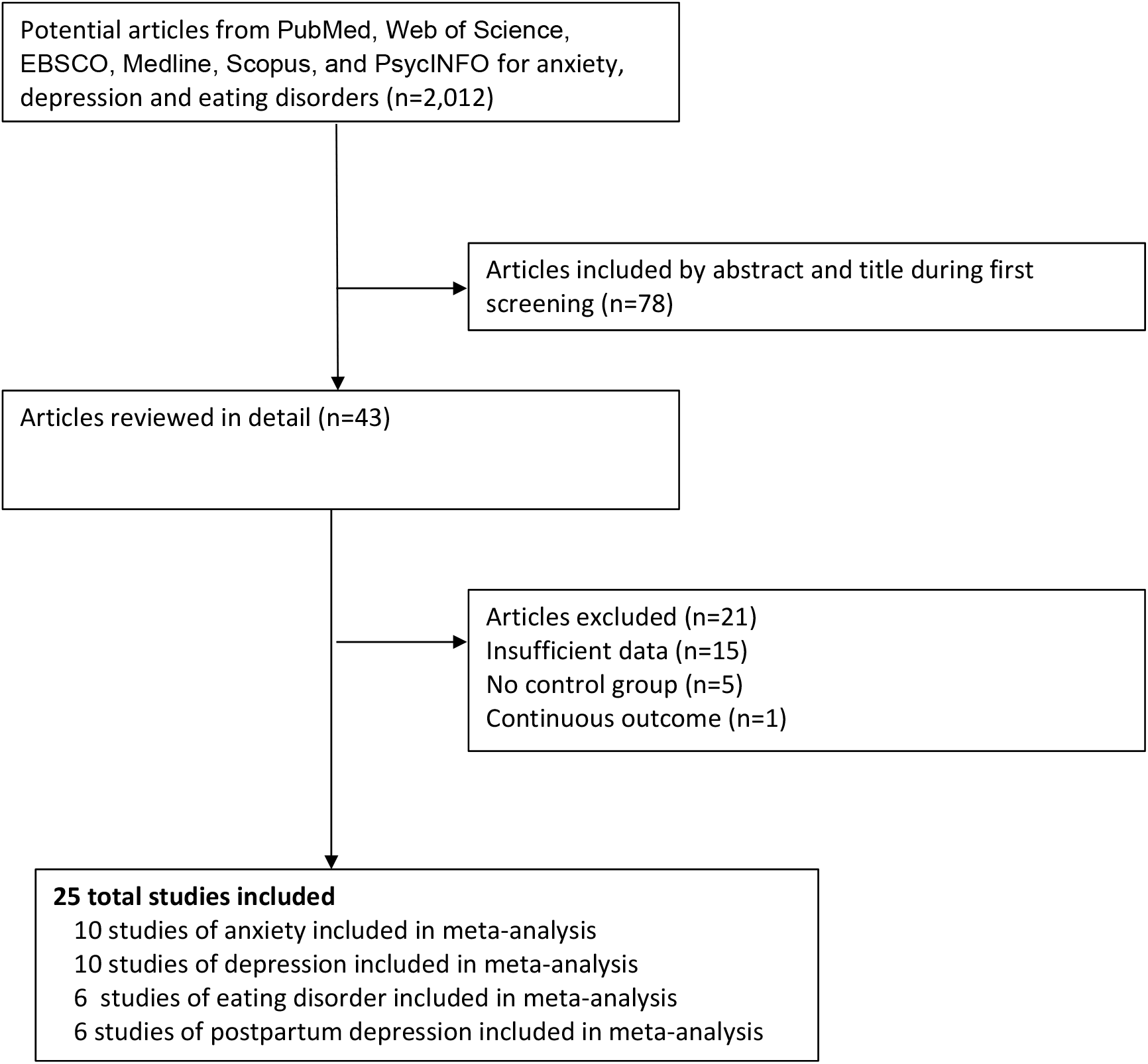
Flow diagram of the literature search and study selection process.

### Excess Prevalence of Anxiety in Women with PCOS

Twelve studies, all assessed as ‘high quality’ (**Supplemental Table 6**), were initially identified measuring as association between anxiety and PCOS (12-23). However, only ten studies were included in the meta-analysis, as two studies were excluded due to the lack of prevalence data (21,22). Compared to age matched women without PCOS, those with PCOS had a higher prevalence of anxiety in the meta-analysis (random effects PR: 1.42; 95% CI: 1.32, 1.52; I^2 43.82%; **Fig. 2A**). For our economic burden calculations, we considered PCOS patients as having a 1.42-fold greater risk of anxiety compared to those without PCOS.

**Fig. 2.**
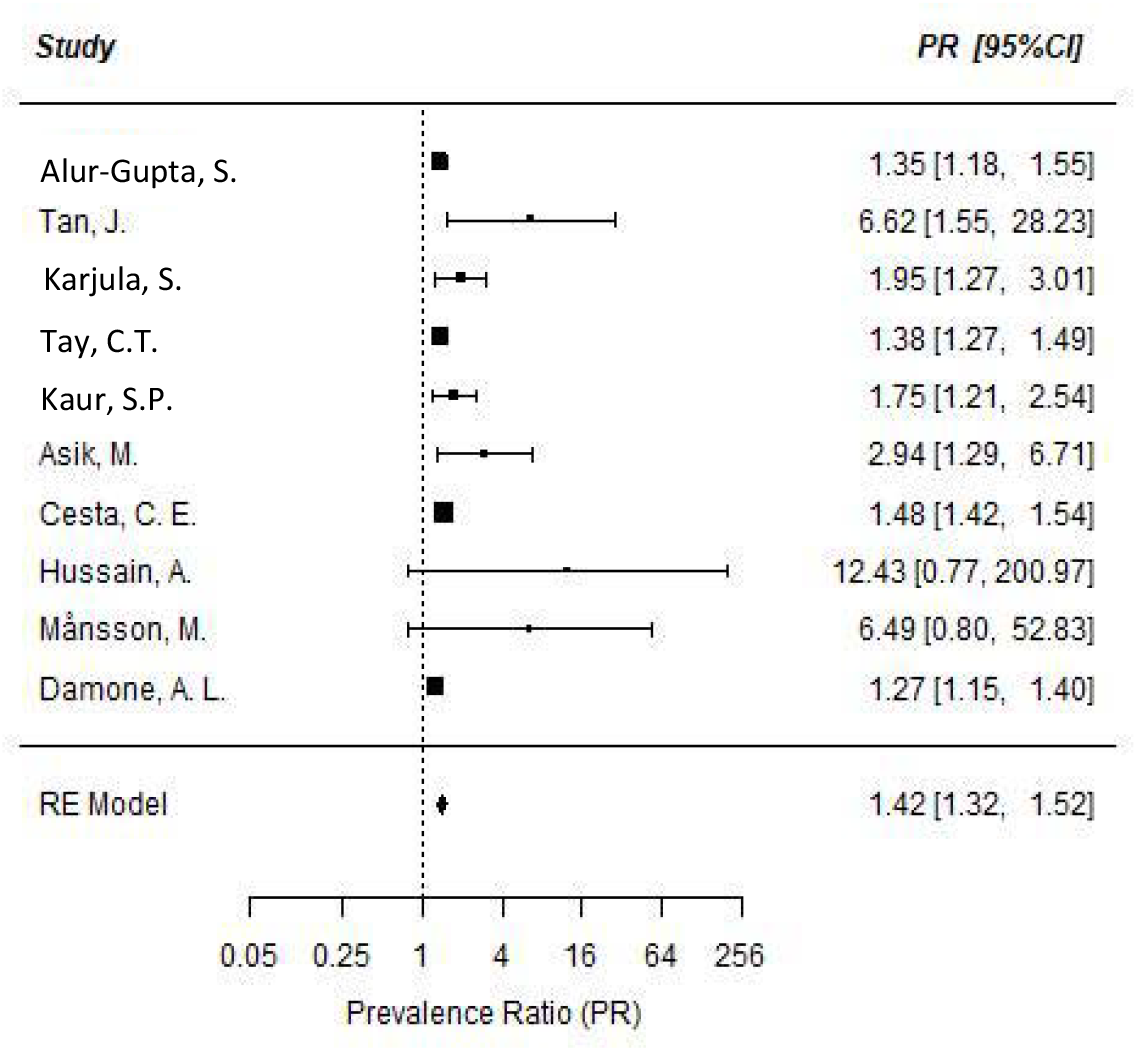
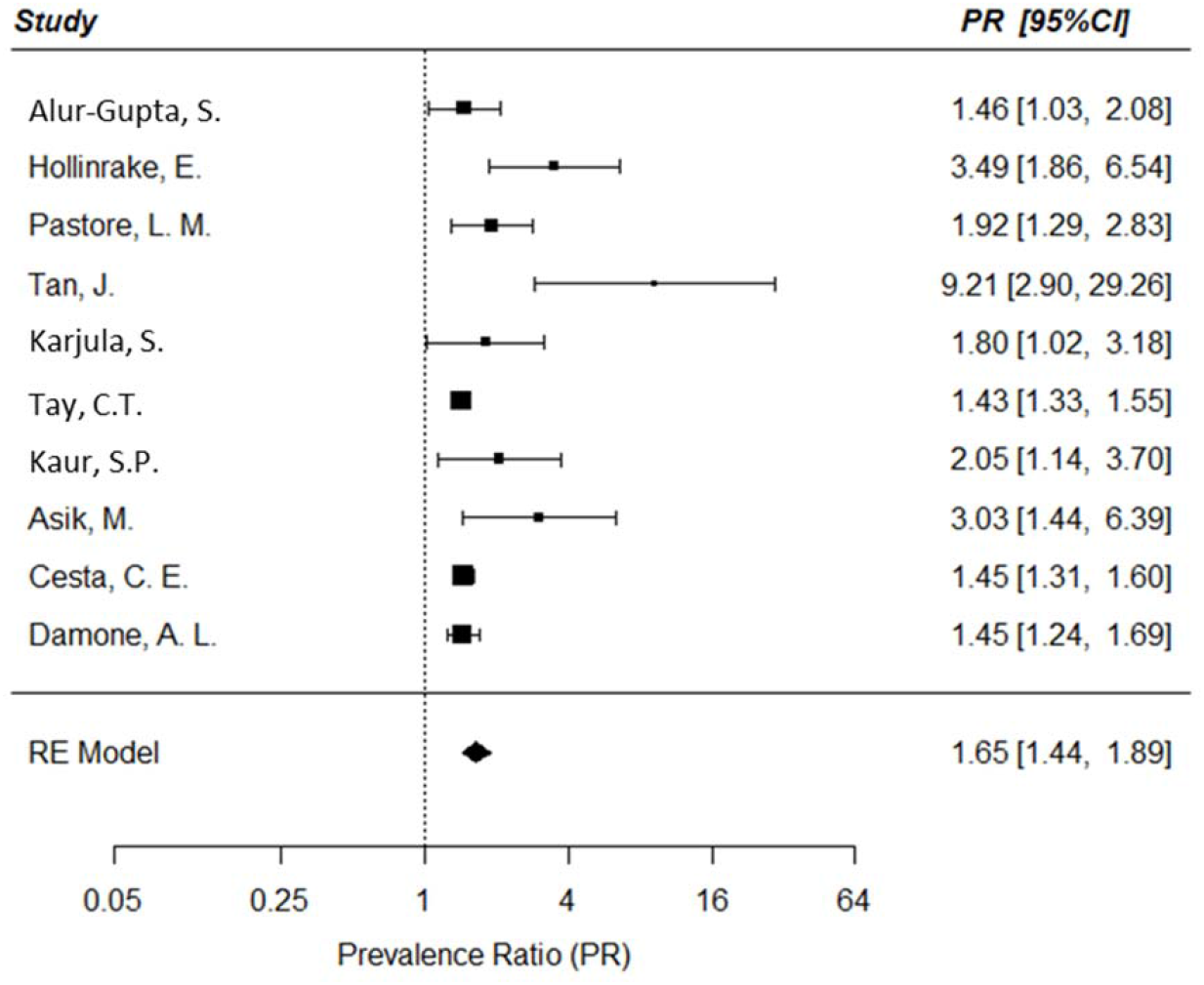
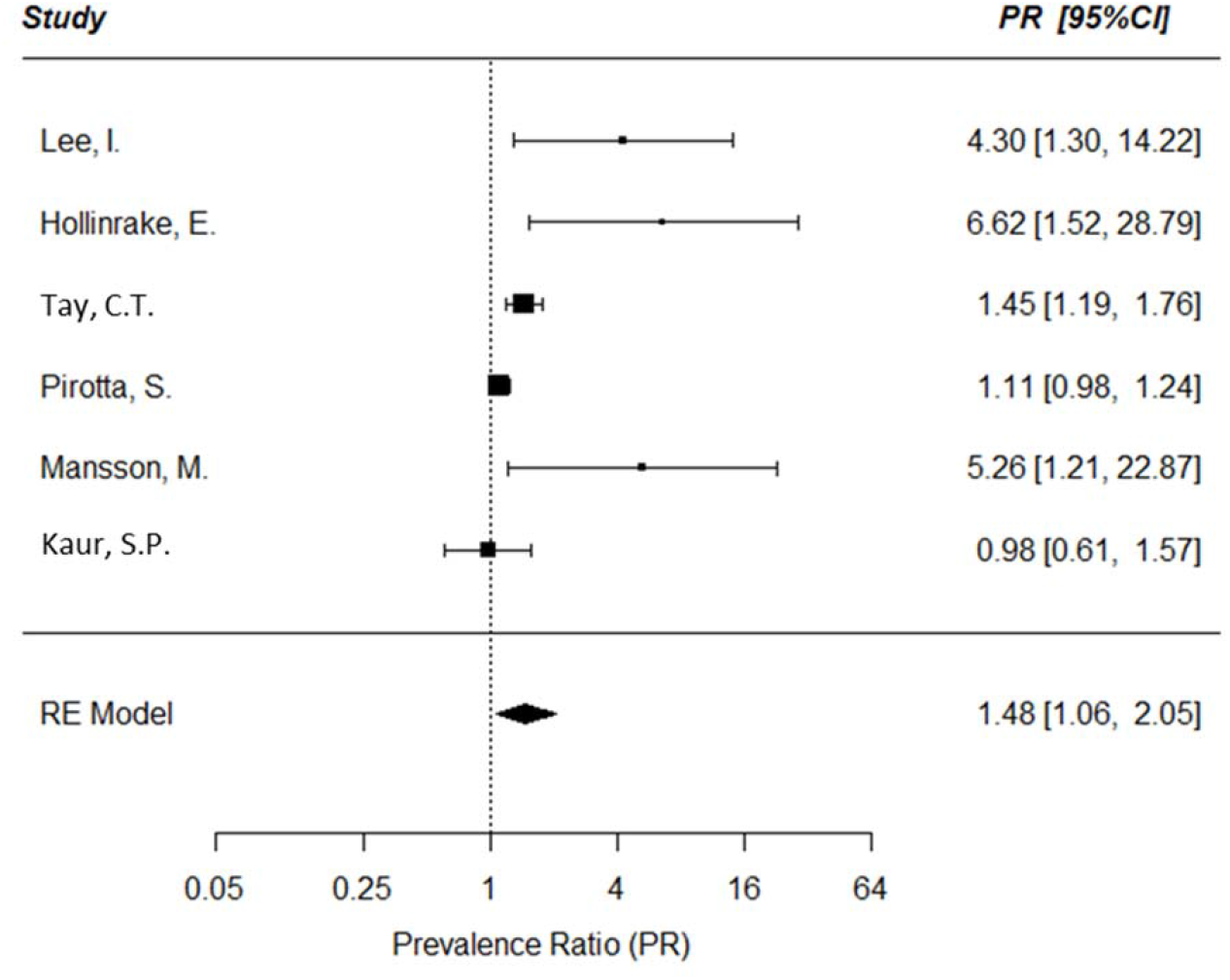
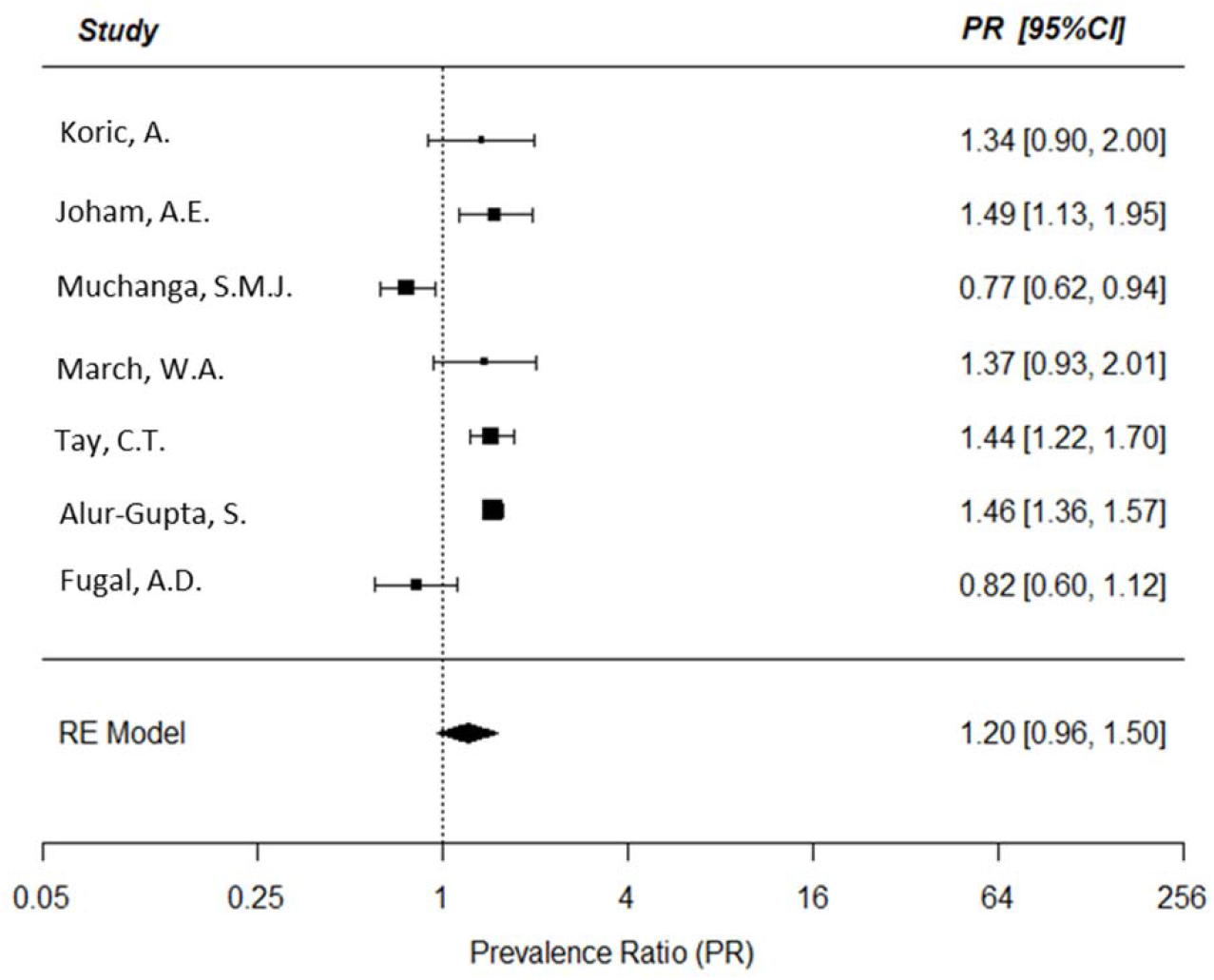
Meta-analyses of the prevalence of mental health disorders in women with PCOS. Forest plots (random effects model) of risk of mental health disorders in women with PCOS, including anxiety (Fig. 2A), depression (Fig. 2B), eating disorders (Fig. 2C), and postpartum depression (Fig. 2D.). See text for abbreviations. **Note:** Studies by Jedel et al. (20) and Li et al. (21) were excluded from the meta-analysis due to the lack of prevalence data.

Considering that comparably aged women of the general population have a prevalence of anxiety of 9.15%, the overall prevalence of anxiety in women with PCOS can be estimated to be 1.42 × 9.15% = 12.99%. The excess prevalence of anxiety due to PCOS is therefore 12.99%-9.15% = 3.84%; the excess number of anxiety cases due to PCOS is 5,631,459 × 3.84% = 216,248 individuals.

### Excess Prevalence of Depression in Women with PCOS

While fifteen studies were initially identified (12-20, 23-27), we used only the ten studies assessed as being of ‘high quality’ (**Supplemental Table 6**) for the meta-analysis. Compared to age matched women without PCOS, those with PCOS had a higher prevalence of depression in the meta-analysis (random effects PR: 1.65; 95% CI: 1.44, 1.89; I^2 63.0%; **Fig. 2B**). For our economic burden calculations, we considered PCOS patients as having a 1.65-fold greater risk of depression compared to those without PCOS.

Considering that comparably aged women of the general population have a prevalence of depression of 8.9%, the overall prevalence of depression in women with PCOS can be estimated to be 1.65 × 8.9% = 14.69%. The excess prevalence of depression due to PCOS is therefore 14.69%-8.9% = 5.79%; the excess number of depression cases due to PCOS is 4,528,088 × 5.79% = 262,176 individuals.

### Excess Prevalence of Eating Disorders in Women with PCOS

Six studies, all assessed as ‘high quality’ (**Supplemental Table 6**), were included for an association between eating disorders and PCOS (14,15,19,23,27,28). Compared to age matched women without PCOS, those with PCOS had a higher prevalence of eating disorders in the meta-analysis (random effects PR: 1.48; 95% CI: 1.06, 2.05; I^2 74.41%; **Fig. 2C**).

Considering that comparably aged women of the general population have a prevalence of eating disorders of 2.4%, the overall prevalence of eating disorders in women with PCOS can be estimated to be 1.48 × 2.4% = 3.55%. The excess prevalence of depression due to PCOS is therefore 3.55% - 2.4% = 1.15%. Therefore, the excess number of anxiety due to PCOS is 4,240,306 × 1.15% = 48,764 individuals.

### Excess Prevalence of Postpartum Depression in Women with PCOS

Six studies, all of high quality (**Supplemental Table 6**), were included examining the association between postpartum depression and PCOS (30-36). Compared to age-matched women without PCOS, those with PCOS had a higher prevalence of postpartum depression in the meta-analysis, however this association did not reach statistical significance (random effects PR: 1.20; 95% CI: 0.96, 1.50; **Fig. 2D**). Because the association between PCOS and postpartum depression was not found to be statistically significant in this meta-analysis, postpartum depression was excluded from our calculations of economic burden.

### Economic Burden of Anxiety in Women with PCOS

The cost of anxiety-related care per individual in need was estimated to be $2,694 in 1990, which converts to $8,966 in 2021 USD. Therefore, the excess cost of anxiety related care in PCOS is 216,248 x $8,966= $1,938,879,568 USD in 2021 (**Table 1**).

**Table 1.** ESTIMATES OF THE EXCESS PREVALENCE AND ECONOMIC BURDEN ASSOCIATED WITH MENTAL HEALTH (MH) MORBIDITIES OF PCOS AS OF 2021 IN THE UNITED STATES.

| **MH Morbidities** | **Excess Prevalence of**<br>**Morbidity in PCOS**<br>**(%)** | **Annual costs in billions in 2021**<br>**USD (% of total costs in**<br>**category)** |
| --- | --- | --- |
| Anxiety | 3.84% | \$1.939 (45.5%) |
| Depression | 5.79% | \$1.678 (39.4%) |
| Eating disorders | 1.15% | \$0.644 (15.1%) |
| **Total excess cost of MH disorders in PCOS** |  | 4.261 (100%) |

### Economic Burden of Depression in Women with PCOS

The cost of depression-related care per individual in need was estimated to be $5,726 in 2018, which converts to $6,401 in 2021 USD. Therefore, the excess cost of depression related care in PCOS is 262,176 x $6,401= $1,678,188,576 USD in 2021 (**Table 1**).

### Economic Burden of Eating Disorders in Women with PCOS

The cost of eating disorder-related care per individual in need was estimated to be $11,808 in 2018, which converts to $13,200 in 2021 USD. Therefore, the excess cost of eating disorder-related care in PCOS is 48,764 x $13,200 = $643,684,800 USD in 2021 (**Table 1**).

### Analyzing for Potential Publication Bias

To assess for possible publication bias, we recalculated the results using the ‘trim and fill’ method. In the “trim and fill” analysis for PCOS-related anxiety the estimated number of missing studies was 4 and the corresponding pooled random effects PR estimate was 1.40 (95% CI: 1.31, 1.50, p<0.001) (**Supplemental Fig. 1A**). In the “trim and fill” analysis for PCOS-related depression the estimated number of missing studies was 4 and the corresponding pooled random effects PR estimate was 1.50 (95% CI: 1.28, 1.76, p<0.0001) (**Supplemental Fig. 1B**). In the “trim and fill” analysis for PCOS-related eating disorders the estimated number of missing studies was 2 and the corresponding pooled random effects PR estimate was 1.30 (95% CI: 0.92, 1.85; p=0.1371) (**Supplemental Fig. 1C**). We did not analyze the data for PCOS-related postpartum depression using the ‘trim and fill’ approach as these results were already not significant.

That the results for PCOS-related eating disorders are no longer significant after applying the ‘trim and fill’ adjustment means that these results were sensitive to one type of selection bias that is due “small study” effects, i.e., the tendency of small studies to suppress publication of results that are negative. However, we should note that the ‘trim and fill’ method cannot be used as formal proof for the presence of publication bias due to “small study” effects, as it is possible that there are other explanations for the lack of symmetry on the funnel plots, including heterogeneity of study populations, covariates, or outcome definitions that may give rise to a lack of symmetry. However, based on this analysis it appears that the results for this outcome are not as robust as for the other outcomes.

## DISCUSSION

In the U.S, the additional direct healthcare costs associated with MH disorders in PCOS were estimated to be $1.939 billion/yr, $1.678 billion/yr., and $0.644 in 2021 USD for anxiety, depression, and eating disorders, respectively. The combined additional direct healthcare costs associated with depression and anxiety in PCOS was estimated to be $4.261 billion/year in 2021 USD, of which 45% can be attributable to anxiety, 40% to depression, and the remainder to eating disorders. While the prevalence of postpartum depression appeared to be increased in PCOS, the difference did not reach significance on meta-analysis and this outcome was not included in our economic burden calculations.

Taken together, including our prior economic burden assessments (2,3), the total excess economic burden estimated for PCOS exceeds $15 billion annually in 2021 USD (**Table 2**). Of this cost, approximately 28% will be accounted for the cost of treating PCOS-related MH disorders, including anxiety, depression and eating disorders; 29.5% will be accounted for the cost of treating reproductive endocrine morbidities (menstrual dysfunction/abnormal uterine bleeding, hirsutism, and infertility); 15.1% is attributable to obstetrical and pregnancy related disorders; and 10.1% and 16.1% is attributable to T2DM and strokes, respectively. The cost of the initial diagnostic evaluation of PCOS is very low ($166 million annually in 2021 USD; **Table 2**), accounting for only 1.1% of the total economic burden attributable to direct healthcare costs of the disorder estimated so far. These data strongly suggest that ensuring quality diagnosis and evaluation for all patients with PCOS is a cost-effective approach to ameliorating the complications and costs associated with the disorder.

**Table 2.**
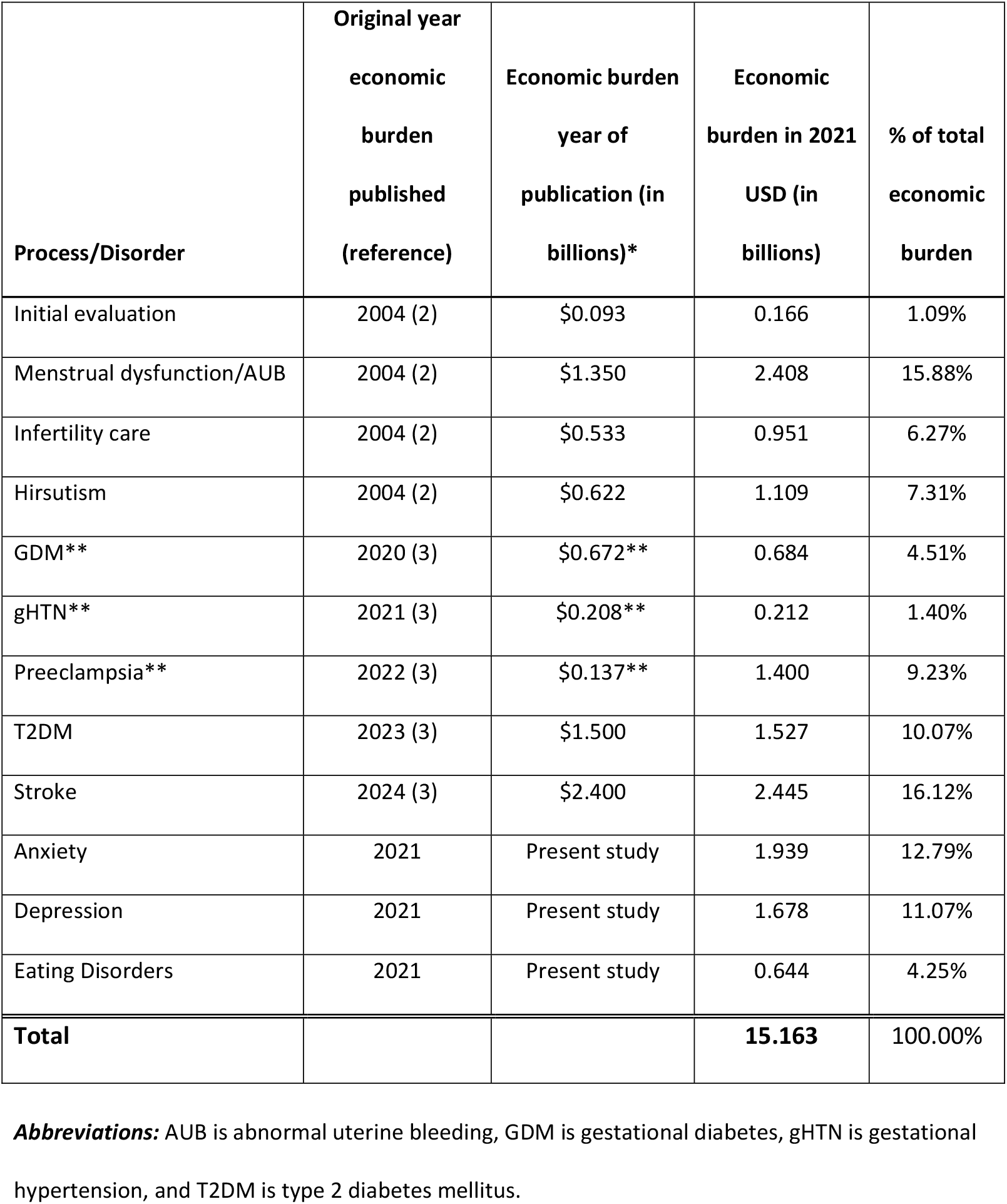
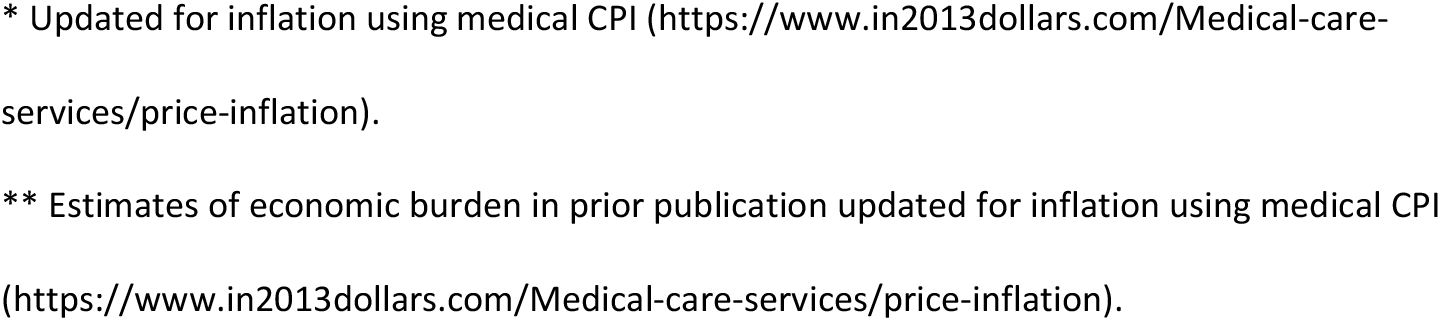
DIRECT HEALTHCARE-RELATED ECONOMIC BURDEN IN PCOS AS OF 2021 IN THE UNITED STATES.

| **Process/Disorder** | **Original year**<br>**economic**<br>**burden**<br>**published**<br>**(reference)** | **Economic burden**<br>**year of**<br>**publication (in**<br>**billions)\*** | **Economic**<br>**burden in 2021**<br>**USD (in**<br>**billions)** | **% of total**<br>**economic**<br>**burden** |
| --- | --- | --- | --- | --- |
| Initial evaluation | 2004 (2) | \$0.093 | 0.166 | 1.09% |
| Menstrual dysfunction/AUB | 2004 (2) | \$1.350 | 2.408 | 15.88% |
| Infertility care | 2004 (2) | \$0.533 | 0.951 | 6.27% |
| Hirsutism | 2004 (2) | \$0.622 | 1.109 | 7.31% |
| GDM** | 2020 (3) | \$0.672** | 0.684 | 4.51% |
| gHTN** | 2021 (3) | \$0.208** | 0.212 | 1.40% |
| Preeclampsia** | 2022 (3) | \$0.137** | 1.400 | 9.23% |
| T2DM | 2023 (3) | \$1.500 | 1.527 | 10.07% |
| Stroke | 2024 (3) | \$2.400 | 2.445 | 16.12% |
| Anxiety | 2021 | Present study | 1.939 | 12.79% |
| Depression | 2021 | Present study | 1.678 | 11.07% |
| Eating Disorders | 2021 | Present study | 0.644 | 4.25% |
| **Total** |  |  | **15.163** | **100.00%** |

**Abbreviations:** AUB is abnormal uterine bleeding, GDM is gestational diabetes, gHTN is gestational hypertension, and T2DM is type 2 diabetes mellitus.
\* Updated for inflation using medical CPI (https://www.in2013dollars.com/Medical-care-services/price-inflation).
\*\* Estimates of economic burden in prior publication updated for inflation using medical CPI (https://www.in2013dollars.com/Medical-care-services/price-inflation).

For perspective, the estimated direct healthcare costs attributable to ovarian cancer, lung cancer, prostate cancer, and breast cancer, in the U.S. are $7.8, $16.8, $26.7, and $20.5 billion in 2021 USD, respectively (37), compared to $15.2 billion for PCOS so far. Furthermore, the included costs are only for those morbidities that to date have been confirmed as increased in PCOS relative to controls after careful meta-analyses considering the quality of the studies. As further studies are undertaken it is likely that the economic burden of PCOS related to direct healthcare costs will continue to rise.

Liu and colleagues assessed the current burden of PCOS at the global, regional, and country-specific levels in 194 countries and territories according to age and socio-demographic index (SDI) (38). The investigators used data from the Global Burden of Diseases, Injuries and Risk Factors Study (GBD) 2017 to estimate the total and age-standard PCOS incidence rates and the associated disability-adjusted life-years (DALYs) rates among women of reproductive age in both 2007 and 2017, and the trends in these parameters from 2007 to 2017. The data sources used in GBD take many forms, including census data, vital registrations, disease registries, survey data, and published and unpublished scientific literature, among other sources (https://www.healthdata.org/acting-data/what-data-sources-go-gbd). These investigators concluded that PCOS accounted for 0.43 million associated DALYs. They also noted slight increases in the age-standardized incidence of PCOS and DALYs among women of reproductive age (15–49 years) from 2007 to 2017 at the global level, and in most regions and countries. Safiri and colleagues also used the GDB Study 2017 database to determine the global, regional, and national burden of PCOS, by age and sociodemographic index (SDI), over the period 1990–2019 (39). These investigators reported that in 2019 the global age-standardized point prevalence and annual incidence rates for PCOS were 1677 (1.7%) and 59 (0.06%) per 100,000, respectively.

Neither one of these studies estimated the economic cost of the burden observed. Furthermore, we should note that Liu et al. (38) reported only on global age-standardized PCOS incidence rates (i.e., the occurrence of *<u>new cases</u>* of PCOS over a specified period of time), not prevalence rates (i.e., the total proportion of persons in the population who have PCOS at a specified point in time or over a specified period of time), among women of reproductive age. While Safiri and colleagues (39) present an estimate of prevalence, we should note that the estimate is significantly lower than that reported when populations are assessed directly for the prevalence of PCOS (1.7% vs. 6.6% or greater), likely reflecting the chronic underdiagnosis of PCOS.

Ding and colleagues estimated the burden of disease attributable to Type 2 DM in women with PCOS using individual patient data from a UK primary care database between 2004 and 2014 and aggregate data from the literature to obtain conversion rates through disease progression (40). A simulation approach was applied to model the population dynamics of PCOS over a follow-up period of 25 years in using Bayesian modeling. The investigators estimated that the associated annual healthcare burden of T2DM in PCOS was at least £237 million in 2014 pounds in the UK. Taking into account the relative populations of reproductive aged women (15-49 years) of the UK and the U.S. (14.7 million vs. 76.5 million) (41), the healthcare inflation rate since 2014 (19.2%), and the conversion rate of pounds to dollars in mid-2021 (0.72 USD to 1 pound), the economic burden for PCOS-associated T2DM estimated by Ding et al is equivalent to $2.04 billion 2021 USD, somewhat higher than the $1.527 2021 USD that we previously estimated (**Table 2**) (3).

Inclusion of only peer-reviewed and controlled studies is a strength of this study. Alternatively, our analysis was limited by the number of studies that met inclusion criteria. This could be due to the fact that PCOS diagnosis creates an umbrella effect that encompasses physical as well as mental disease which creates a false belief in patients that they cannot be treated or diagnosed with a disease other than PCOS due to the misconception that their symptoms stem solely from the PCOS diagnosis. Other limitations of this study include the fact that while the cost estimates were conducted with for the U.S., the meta-analysis used some studies from other countries. Finally, while other MH disorders have been shown to be associated with PCOS, such as bipolar disorder and obsessive-compulsive disorder, we focused our study on the four most common disorders found in the literature (4,5).

Overall, our current study suggests that the additional direct healthcare costs due to PCOS-related anxiety, depression and eating disorders exceeds $4 billion annually in 2021 USD in the United States. So far, the total direct economic burden of PCOS exceeds $15 billion annually in 2021 USD, and MH disorders account for almost one-third of these costs. Notably, these estimates are solely for the United States and PCOS is a global disease. As PCOS is clinically most apparent during the reproductive age, we should note that the number of women between the ages of 15 and 49 worldwide is estimated to be about 1.9 billion (41). Taking into account that most studies assessing the prevalence of PCOS report a minimum rate similar to what was used on the present study (i.e., 6.6% based on the NIH criteria) (42), we can estimate that there are *<u>at least</u>* 125.4 million women affected with PCOS worldwide. Consequently, the world-wide economic burden for PCOS can be assumed to be enormous.

While we have examined the direct economic burden related to the medical or health-related costs of the disorder, we should note that a complete understanding of the economic burden of the disorder requires that we also assess the indirect (those attributable to loss of work productivity) and intangible (those related to the pain and sufferings of patients because of a disorder, usually measured by using the reduction in quality of life) costs of the disorder. In conclusion, these data suggest that improved detection of PCOS and more significant clinical awareness and interventions for PCOS and its associated disorders is a cost-effective approach to ameliorating the economic, health, and quality of life impact of PCOS. Finally, our findings further highlight the need to increase investment in PCOS research, which is currently severely underfunded relative to the economic burden of the disorder (43).

## Data Availability

Surabhi Yadav, Olivia Delau, Adam Bonner, Daniela Markovic, William Patterson, Sasha Ottey, and Richard P. Buyalos, have no competing interests to declare. Ricardo Azziz serves as a consultant for Spruce Bioscience, Core Access Surgical Technology, Rani Therapeutics, and Fortress Biotech, advisor for Arora Forge, and investor in Martin Imaging.

---

**Supplementary Table 1.** Search terms used for systematic review.

| Topic | Keywords |
| --- | --- |
| Mental Disorders in PCOS | "Polycystic Ovary Syndrome/psychology"[MAJR] |
|  | 'polycystic ovary syndrome' AND 'mental disorder' OR 'depression' OR 'anxiety' |
|  | 'polycystic ovary syndrome' AND 'mental illness' |
|  | ("polycystic ovary syndrome" AND "mental disorder") AND (LIMIT-TO (LANGUAGE, "English")) |
| Direct costs for Mental Disorders in PCOS | "PCOS" AND "economic burden" OR "costs" OR "cost-of-illness" OR "burden of illness" AND "mental health" |
|  | "PCOS" AND "economic burden" AND "mental health" |
| Economic burden of Mental Disorders | (mental health OR mental illness OR mental disorder OR psych* OR anxiety OR depression OR quality of life OR bulimia OR bipolar disorder) AND ('cost*' OR 'economic burden' OR 'cost-of-illness' OR 'burden of illness') |
|  | "Depressive Disorder/economics"[MAJR] |
|  | "Depressive Disorder/economics"[MAJR] |

**Supplementary Table 2.** Characteristics of Included Studies Categorized by Anxiety.

| **Study** | **Year of Publication** | **Country** | **Study Design** | **Total Sample Size per Group** | **PCOS Definition** |
| --- | --- | --- | --- | --- | --- |
| Alur-Gupta et al.<sup>11</sup> | 2019 | US | Case-Control | PCOS= 189<br>Controls= 225 | Rotterdam Criteria |
| Tan et al.<sup>12</sup> | 2017 | China | Case-Control | PCOS= 120<br>Controls= 100 | Rotterdam Criteria |
| Karjula et al.<sup>13</sup> | 2017 | Finland | Population-based Cohort Study | PCOS= 125<br>Controls= 2188 | Self-reported PCOS |
| Tay et al.<sup>14</sup> | 2020 | Australia | Cross-Sectional | PCOS=760<br>Controls= 7910 | Self-reported PCOS |
| Kaur et al.<sup>15</sup> | 2019 | India | Cross-Sectional | PCOS= 73<br>Controls= 78 | Rotterdam Criteria |
| Asik et al.<sup>16</sup> | 2015 | Germany | Cross-sectional | PCOS=71<br>Controls= 50 | Rotterdam Criteria |
| Cesta et al.<sup>17</sup> | 2016 | Sweden | Matched Cohort Design | PCOS= 24385<br>Controls= 243850 | ICD-8, ICD-9, ICD-10 codes |
| Hussain et al.<sup>18</sup> | 2015 | India | Case-Control | PCOS=110<br>Controls=40 | Diagnosis of PCOS was confirmed according to the National Institute of Health/National Institute of Child Health and Human Development, 1990 consensus conference criteria |
| Mansson et al.<sup>19</sup> | 2008 | Sweden | Case-Control | PCOS= 49<br>Controls= 49 | Rotterdam Criteria |
| Jedel et al.<sup>20</sup> | 2009 | Sweden | Case-Control | PCOS=30<br>Controls=30 | 12 or more 2–9 mm ovarian follicles and/or ovarian volume exceeding 10 ml in one or two ovaries; clinical signs of hyperandrogenism and/or oligo/amenorrhea. |
| Li et al.<sup>21</sup> | 2017 | China | observational | PCOS= 103<br>Controls=110 | Rotterdam Criteria |
| Damone et al.<sup>22</sup> | 2018 | Australia | cross sectional analysis | PCOS=478<br>Controls=8134 | Self-reported PCOS |

**Supplementary Table 3.** Characteristics of Included Studies Categorized by Depression.

| Study | Year | Country | Study Design | Total Sample Size per Group | PCOS Definition |
| --- | --- | --- | --- | --- | --- |
| Alur-Gupta et al.<sup>11</sup> | 2019 | US | Cross-sectional | PCOS= 189<br>Controls= 225 | Rotterdam criteria |
| Hollinrake et al.<sup>23</sup> | 2006 | US | Prospective longitudinal study | PCOS= 103<br>Controls= 103 | Rotterdam criteria |
| Pastore et al.<sup>24</sup> | 2004 | US | Cross-sectional | PCOS= 94<br>Controls= 96 | Diagnosis of PCOS, as confirmed through the study using the NICHD criteria of oligomenorrheic and non-diabetic, with self-reported hirsutism and/or acne and/or elevated free testosterone (>6.8 pg/ml) |
| Tan et al.<sup>12</sup> | 2017 | China | Case-control | PCOS= 120<br>Controls= 100 | Rotterdam criteria |
| Karjula et al.<sup>13</sup> | 2017 | Finland | Population-based follow-up | PCOS= 125<br>Controls= 2188 | Self-reported PCOS |
| Tay et al.<sup>14</sup> | 2020 | Australia | Cross sectional study | PCOS=760<br>Controls= 7910 | Self-reported PCOS |
| Kaur et al.<sup>15</sup> | 2019 | India | Cross sectional case control study | PCOS= 73<br>Controls= 78 | Rotterdam criteria |
| Asik et al.<sup>16</sup> | 2015 | Germany | Cross-sectional | PCOS=71<br>Controls=50 | Rotterdam criteria |
| Cesta et al.<sup>17</sup> | 2016 | Sweden | Matched cohort design | PCOS= 24385<br>Controls= 243850 | ICD-8, ICD-9, ICD-10 codes |
| Cinar et al.<sup>25</sup> | 2011 | Turkey | Case control | PCOS= 226<br>Controls=85 | Rotterdam criteria |
| Hussain et al.<sup>18</sup> | 2015 | India | Case control | PCOS=110 | Diagnosis of PCOS was confirmed |
|  |  |  |  | Controls=40 | according to the National Institute of Health/National Institute of Child Health and Human Development, 1990 consensus conference criteria |
| Mansson et al.<sup>19</sup> | 2008 | Sweden | Case-control study | PCOS= 49<br>Controls= 49 | Rotterdam criteria |
| Adali et al.<sup>26</sup> | 2008 | Turkey | Case control study | PCOS=42<br>Controls=42 | Positive diagnosis is based on the patient presenting any two of the following three features: oligo- or anovulation, clinical and/or biochemical signs of hyperandrogenism, and polycystic ovaries on ultrasound examination |
| Damone et al.<sup>22</sup> | 2018 | Australia | Cross-sectional | PCOS=478<br>Controls=8134 | Self-reported PCOS |
| Jedel et al.<sup>20</sup> | 2010 | Sweden | Case control | PCOS=30<br>Controls=30 | 12 or more 2–9 mm ovarian follicles and/or ovarian volume exceeding 10 ml in one or two ovaries; clinical signs of hyperandrogenism and/or oligo/amenorrhea. |

**Supplementary Table 4.** Characteristics of Included Studies Categorized by Eating Disorders.

| Study | Year | Country | Study Design | Total Sample Size per Group | PCOS Definition |
| --- | --- | --- | --- | --- | --- |
| Lee et al.<sup>27</sup> | 2015-16 | US | Cross-sectional study | PCOS = 148<br>Controls = 106 | Rotterdam criteria |
| Hollinrake et al.<sup>23</sup> | 2006 | US | Prospective Cohort Study | PCOS= 103<br>Controls= 103 | Rotterdam Criteria |
| Tay et al.<sup>14</sup> | 2020 | Australia | Cross-Sectional | PCOS= 875<br>Controls= 7592 | Self-reported PCOS |
| Pirotta et al.<sup>28</sup> | 2019 | Australia | Cross-Sectional | PCOS= 501<br>Controls= 398 | Self-reported PCOS |
| Kaur et al.<sup>15</sup> | 2019 | India | Cross-Sectional | PCOS= 73<br>Controls= 78 | Rotterdam Criteria |
| Mansson et al.<sup>19</sup> | 2008 | Sweden | Case-control | PCOS= 49<br>Controls= 49 | Rotterdam Criteria |

**Supplementary Table 5.** Characteristics of Included Studies Categorized by Postpartum Depression.

| Study | Year | Country | Study Design | Total Sample Size per Group | PCOS Definition |
| --- | --- | --- | --- | --- | --- |
| Koric et al.<sup>29</sup> | 2021 | US | Cases= 320<br>Controls= 3586 | PCOS= 320<br>Controls= 3586 | “Have you ever been told that you have Polycystic Ovarian Syndrome or PCOS by a doctor, nurse, or other healthcare worker?”<br>PCOS symptomatology was defined in possible alternate ways as having (1) irregular periods and acne, (2) irregular periods and hirsutism, or (3) irregular periods, acne, and hirsutism. |
| Joham et al.<sup>30</sup> | 2016 | Australia | Cross-Sectional | PCOS= 320<br>Controls= 4578 | Self-reported PCOS |
| Muchanga et al.<sup>31</sup> | 2017 | Japan | Population-based Prospective Birth Cohort Study | Cases with postpartum depression= 11,341<br>Controls without postpartum depression= 71,148 | Self-reported PCOS |
| March et al.<sup>32</sup> | 2018 | Australia | Cross-Sectional | PCOS= 52<br>Controls= 514 | Rotterdam Criteria |
| Tay et al.<sup>33</sup> | 2019 | Australia | Cross-Sectional | PCOS= 436<br>Controls= 4803 | Self-reported PCOS |
| Alur-Gupta et al.<sup>34</sup> | 2021 | US | Retrospective Cohort Study | PCOS= 42,391<br>Controls= 795,480 | ICD-9, ICD-10 |
| Fugal et al.<sup>35</sup> | 2022 | US | Population-based Study | PCOS= 3,280<br>Controls= 48,348 | Rotterdam Criteria |

**Supplementary Table 6.** Quality scores of studies using Newcastle-Ottawa Scale.

| Study | Selection |  |  |  | Comparability | Outcome |  |  | NOS |
| --- | --- | --- | --- | --- | --- | --- | --- | --- | --- |
|  | Representative-ness of the exposed cohort | Adequate control selection | Adequate end-point definition | Demonstration that outcomes was not present at start of study | Comparability on the basis of the design or analysis | Assessment of outcome | Adequate follow-up duration | Adequate follow-up rate | Overall score |
| **Anxiety** |  |  |  |  |  |  |  |  |  |
| Alur-Gupta et al.<sup>11</sup> | 1 | 1 | 1 | 1 | 1 | 1 | 1 | 1 | 8 |
| Tan et al.<sup>12</sup> | 1 | 1 | 1 | 1 | 0 | 1 | 1 | 1 | 7 |
| Karjula et al.<sup>13</sup> | 0 | 1 | 1 | 1 | 1 | 1 | 1 | 1 | 7 |
| Tay et al.<sup>14</sup> | 1 | 1 | 1 | 1 | 1 | 1 | 1 | 1 | 8 |
| Kaur et al.<sup>15</sup> | 1 | 1 | 1 | 1 | 0 | 1 | 1 | 1 | 7 |
| Asik et al.<sup>16</sup> | 0 | 1 | 1 | 1 | 1 | 1 | 1 | 1 | 7 |
| Cesta et al.<sup>17</sup> | 1 | 1 | 1 | 1 | 2 | 1 | 0 | 0 | 7 |
| Hussain et al.<sup>18</sup> | 0 | 1 | 1 | 1 | 1 | 1 | 1 | 1 | 7 |
| Mansson et al.<sup>19</sup> | 0 | 1 | 1 | 1 | 1 | 1 | 1 | 1 | 7 |
| Jedel et al.<sup>20</sup> | 1 | 1 | 1 | 1 | 0 | 1 | 1 | 1 | 7 |
| Li et al.<sup>21</sup> | 1 | 1 | 1 | 1 | 1 | 1 | 1 | 1 | 8 |
| Damone et al.<sup>22</sup> | 1 | 1 | 1 | 1 | 1 | 1 | 1 | 0 | 7 |
| **Mean NOS overall score for Anxiety** |  |  |  |  |  |  |  |  | 7.25 |
| **Depression** |  |  |  |  |  |  |  |  |  |
| Alur-Gupta et al.<sup>11</sup> | 1 | 1 | 1 | 1 | 1 | 1 | 1 | 1 | 8 |
| Hollinrake et al.<sup>23</sup> | 1 | 1 | 1 | 1 | 1 | 1 | 1 | 1 | 8 |
| Pastore et al.<sup>24</sup> | 0 | 1 | 1 | 1 | 2 | 1 | 1 | 1 | 8 |
| Tan et al.<sup>12</sup> | 1 | 1 | 1 | 1 | 0 | 1 | 1 | 1 | 7 |
| Karjula et al.<sup>13</sup> | 0 | 1 | 1 | 1 | 1 | 1 | 1 | 1 | 7 |
| Tay et al.<sup>14</sup> | 1 | 1 | 1 | 1 | 1 | 1 | 1 | 1 | 8 |
| Kaur et al.<sup>15</sup> | 1 | 1 | 1 | 1 | 0 | 1 | 1 | 1 | 7 |
| Asik et al.<sup>16</sup> | 0 | 1 | 1 | 1 | 1 | 1 | 1 | 1 | 7 |
| Cesta et al.<sup>17</sup> | 1 | 1 | 1 | 1 | 2 | 1 | 0 | 0 | 7 |
| Cinar et al.<sup>25</sup> | 0 | 0 | 1 | 0 | 0 | 1 | 1 | 1 | 4 |
| Hussain et al.<sup>18</sup> | 0 | 1 | 1 | 0 | 0 | 0 | 0 | 0 | 2 |
| Mansson et al.<sup>19</sup> | 0 | 1 | 1 | 0 | 0 | 1 | 0 | 0 | 3 |
| Adali et al.<sup>26</sup> | 0 | 0 | 1 | 1 | 0 | 1 | 1 | 0 | 4 |
| Damone et al.<sup>22</sup> | 1 | 1 | 1 | 1 | 1 | 1 | 1 | 0 | 7 |
| Jedel et al.<sup>20</sup> | 0 | 1 | 1 | 0 | 0 | 1 | 1 | 1 | 5 |
| **Mean NOS overall score for depression** |  |  |  |  |  |  |  |  | 6.13 |
| **Eating Disorder** |  |  |  |  |  |  |  |  |  |
| Lee et al.<sup>27</sup> | 1 | 1 | 1 | 1 | 1 | 1 | 1 | 1 | 8 |
| Hollinrake et al.<sup>23</sup> | 1 | 1 | 1 | 1 | 1 | 1 | 1 | 1 | 8 |
| Tay et al.<sup>14</sup> | 1 | 1 | 1 | 1 | 1 | 1 | 1 | 1 | 8 |
| Pirotta et al.<sup>28</sup> | 1 | 1 | 1 | 1 | 1 | 1 | 1 | 1 | 8 |
| Kaur et al.<sup>15</sup> | 1 | 1 | 1 | 1 | 0 | 1 | 1 | 1 | 7 |
| Mansson et al.<sup>19</sup> | 0 | 1 | 1 | 1 | 1 | 1 | 1 | 1 | 7 |
| **Mean NOS overall score for eating disorders** |  |  |  |  |  |  |  |  | 7.67 |
| **Postpartum Depression** |  |  |  |  |  |  |  |  |  |
| Koric et al.<sup>29</sup> | 1 | 1 | 1 | 1 | 1 | 1 | 1 | 1 | 8 |
| Joham et al.<sup>30</sup> | 1 | 1 | 1 | 1 | 1 | 1 | 1 | 1 | 8 |
| Muchanga et al.<sup>31</sup> | 1 | 1 | 1 | 1 | 1 | 1 | 1 | 1 | 8 |
| March et al.<sup>32</sup> | 0 | 1 | 1 | 1 | 1 | 1 | 1 | 1 | 7 |
| Tay et al.<sup>33</sup> | 1 | 1 | 1 | 1 | 1 | 1 | 1 | 1 | 8 |
| Alur-Gupta et al.<sup>34</sup> | 1 | 1 | 1 | 1 | 1 | 1 | 1 | 1 | 8 |
| Fugal et al..<sup>35</sup> | 0 | 1 | 1 | 1 | 1 | 1 | 1 | 1 | 7 |
| **Mean NOS overall score for postpartum depression** |  |  |  |  |  |  |  |  | 7.71 |

**Supplemental Figure 1.**
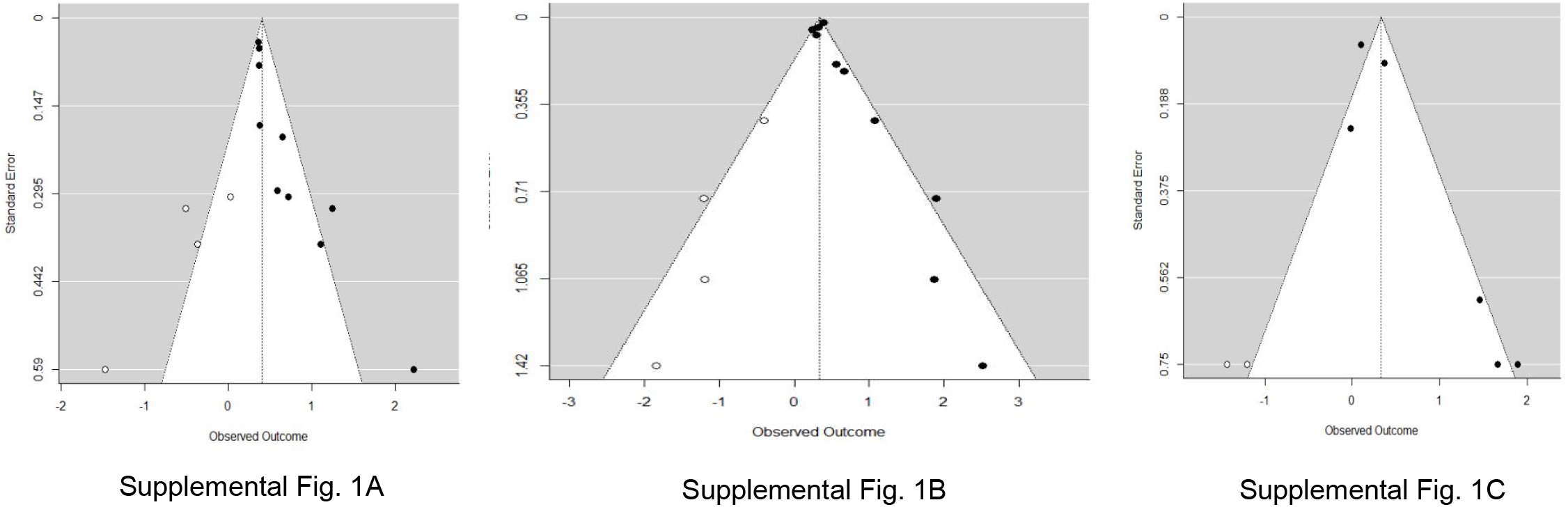
Trim and Fill Funnel Plots for the meta-analyses of anxiety, depression, and eating disorders. Trim and fill funnel plots (X axis is the log prevalence ratio/risk ratio; Y axis is the standard error of the log prevalence ratio/risk ratio) include 10 high quality studies for anxiety (**Supplemental Fig. 1A**), 10 high quality studies for depression (**Supplemental Fig. 1B**), and 6 high quality studies for eating disorders (**Supplemental Fig. 1C**). Observed studies are indicated by solid circles, and “filled” studies are indicated by open circles. The results of the trim and fill analysis indicated that the pooled estimate for anxiety was robust to “small study” effects. The pooled estimate for depression may be slightly overestimated due to the suppression of more negative results on the left side of the funnel plot for smaller studies. It is likely that the over-estimation in the results for depression and eating disorders is caused by publication bias, but this cannot be formally proven. Alternative explanations of funnel plot asymmetry are possible. For example, funnel plot asymmetry can be caused by between study heterogeneity. Different studies may estimate different effects due to differences in study design, characteristics of the study sample, outcome definitions or geographic location. For depression, the removal of one outlier (study by Tan et al [12]) from the meta-analysis eliminated about 30% of the between-study heterogeneity.

